# Leveraging Electronic Medical Records and Knowledge Networks to Predict Disease Onset and Gain Biological Insight Into Alzheimer’s Disease

**DOI:** 10.1101/2023.03.14.23287224

**Authors:** Alice Tang, Katherine P. Rankin, Gabriel Cerono, Silvia Miramontes, Hunter Mills, Jacquelyn Roger, Billy Zeng, Charlotte Nelson, Karthik Soman, Sarah Woldemariam, Yaqiao Li, Albert Lee, Riley Bove, Maria Glymour, Tomiko Oskotsky, Zachary Miller, Isabel Allen, Stephan J. Sanders, Sergio Baranzini, Marina Sirota

## Abstract

Early identification of Alzheimer’s Disease (AD) risk can aid in interventions before disease progression. We demonstrate that electronic health records (EHRs) combined with heterogeneous knowledge networks (e.g., SPOKE) allow for (1) prediction of AD onset and (2) generation of biological hypotheses linking phenotypes with AD. We trained random forest models that predict AD onset with mean AUROC of 0.72 (-7 years) to .81 (-1 day). Top identified conditions from matched cohort trained models include phenotypes with importance across time, early in time, or closer to AD onset. SPOKE networks highlight shared genes between top predictors and AD (e.g., APOE, IL6, TNF, and INS). Survival analysis of top predictors (hyperlipidemia and osteoporosis) in external EHRs validates an increased risk of AD. Genetic colocalization confirms hyperlipidemia and AD association at the APOE locus, and AD with osteoporosis colocalize at a locus close to MS4A6A with a stronger female association.

## Introduction

Neurodegenerative disorders are devastating, heterogeneous, and challenging to diagnose, and their burden in an aging population is expected to continue to grow^1^. Among these, Alzheimer’s Disease (AD) is the most common form of dementia after age 65, and its hallmark memory loss and other cognitive symptoms are costly and onerous to both patients and caregivers. Approaches to curb this impact are moving increasingly to targeting interventions in at-risk individuals prior to the onset of irreversible decline^2–4^. To this end, advancements in AD biomarkers, diagnostic tests, and neuroimaging have improved the detection and classification of AD, and disease-modifying treatments have been approved, but there is still no cure and much remains unknown about its pathogenesis^5,6^. This is in part due to limited availability of longitudinal data or data linking molecular and clinical domains.

In the past few decades, electronic health records (EHRs) have become a source of rich longitudinal data that can be leveraged to understand and predict complex diseases, particularly AD. Prior applications of EHRs for studying AD include deep phenotyping of AD^7^, identification of AD-related associations and hypotheses^8^, and models classifying or predicting a dementia diagnosis from clinical data modalities^9^. Data available in clinical records can also better represent a clinician’s knowledge of a patient’s clinical history at a point in time prior to further diagnostic studies or imaging, allowing a prediction model to be low cost to implement as a first line application in primary care or for initial risk stratification^10^. While machine learning (ML) has been previously applied to EHRs for general dementia classification and prediction^11–13^, these approaches are limited in their specificity for the AD phenotype, lack of biological interpretability, or rely on data modalities that may not be readily available in the EHR to facilitate early prediction (e.g. neuroimaging^14–16^ or special biomarkers^17,18^). Sex as a biological variable is an important covariate for AD heterogeneity with potential contributions to differing risks and resilience, but sex-specific contributions have often been omitted from prior AD machine learning models^19,20^. To our knowledge, there have not yet been approaches that utilize vast EHR data for predicting future risk of AD with consideration of applicability and explainability of models.

With recent advances in informatics and curation of multi-omics knowledge, there is increasing interest in integrative approaches to derive insights into disease. Heterogeneous biological knowledge networks bring in the ability to synthesize decades of research and combine human understanding of multi-level biological relationships across genes, pathways, drugs, and phenotypes, with vast potential for deriving biological meaning from clinical data^21^. There has been much AD-research leveraging specific data modalities or combining a few modalities (transcriptomics^22,23^, genetics^24^, neuroimaging^25^), but there is still a need for meaningful integration that allows for the understanding of the relationship between pathogenesis and clinical manifestations. Heterogeneous knowledge networks provide an opportunity to derive biological hypotheses from clinical data by synthesizing knowledge across multiple data modalities to explain potential relationships between shared clinical associations^26,27^.

In this paper, we utilize EHR data from the University of California, San Francisco (UCSF) medical center to develop ML models for AD onset prediction and generate hypotheses of high-level biological relationships between top predictors and AD. We carry out clinical model construction for prediction and proceed with interpretation of matched patient models, controlling for demographics and visit-related confounding, to identify biologically relevant clinical predictors. We further demonstrate interpretability using heterogeneous knowledge networks (SPOKE knowledge graph)^28^ and validate predictors with supporting evidence in external EHR datasets and through genetic colocalization analysis. Our work not only has implications for determining clinical risk of AD based on EHRs, but also can lead to further research in identifying hypothesized early phenotypes and pathways to help further the field of neurodegeneration.

## Results

From the UCSF EHR database of over 5 million patients, 2,996 AD patients who had undergone dementia evaluation at the Memory and Aging Center and thus had expert-level clinical diagnoses were identified and mapped to the UCSF Observational Medical Outcomes Partnership (OMOP) EHR database. From the remaining patients, 823,671 control patients were extracted with over a year of visits and no dementia diagnosis. After identifying an index time representing AD onset (see Methods) and filtering for availability of at least 7 years of longitudinal data, 749 AD patients and 250,545 control patients were identified (demographics shown in Table 1). From that, 30% was held-out for model evaluation and 70% utilized for model training (Figure 1B). For each time point and within sex strata, ML models were either trained for AD onset prediction or trained on the AD cohort and a subset of propensity-score matched controls for hypothesis generation, where balancing was performed on demographics (sex, race, ethnicity, birth year, prediction age) and visit-related factors (years in EHR, first EHR visit age, number of visits, number of EHR concepts, and days since first EHR record, Supplemental Table 3, matched example in Table 1).

**Table 1:**
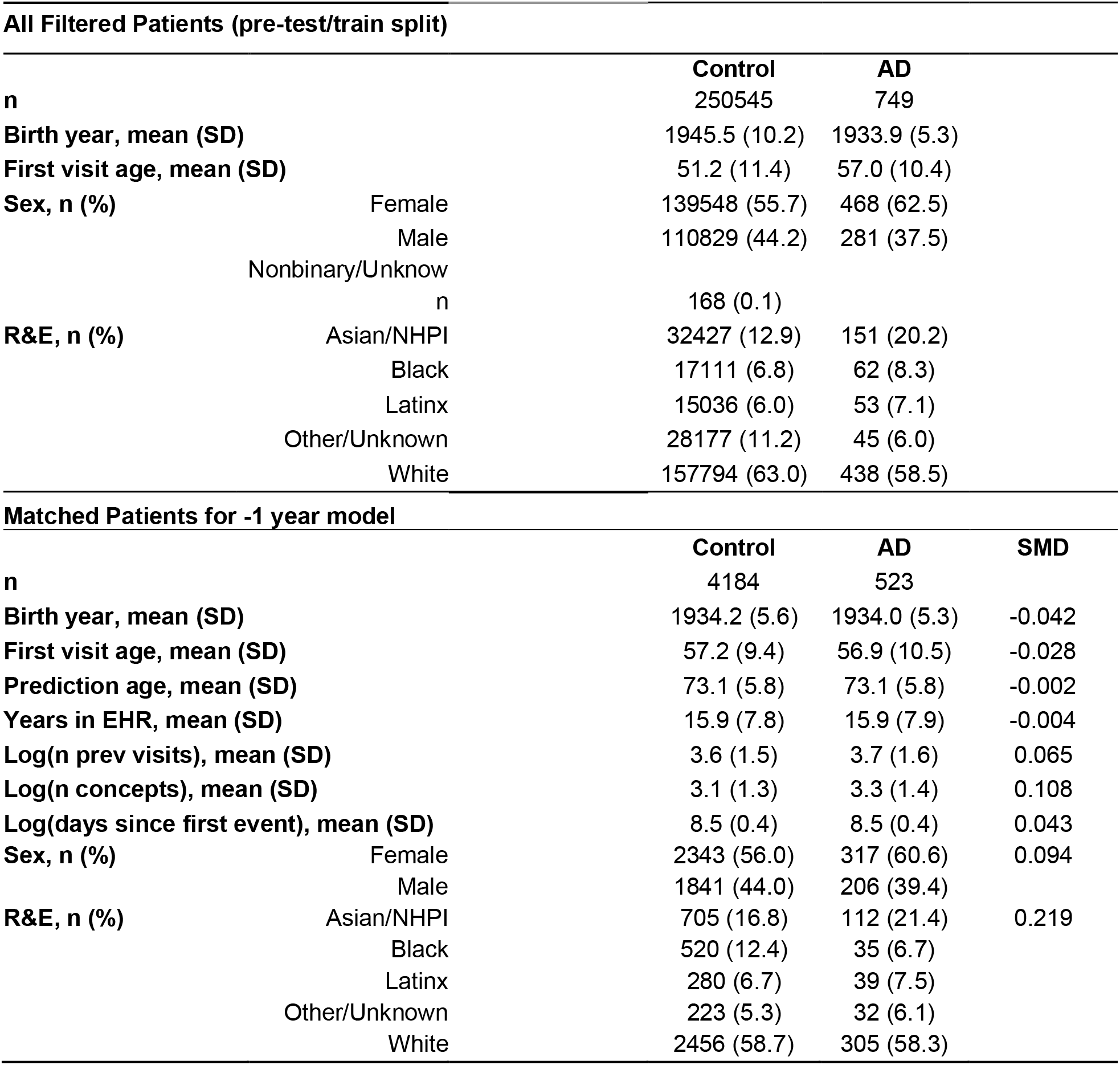
Demographics of patients used in models, and an example matched cohort for the −1 year model

| All Filtered Patients (pre-test/train split) |  |  |  |  |
| --- | --- | --- | --- | --- |
|  |  | Control | AD |  |
| n |  | 250545 | 749 |  |
| Birth year, mean (SD) |  | 1945.5 (10.2) | 1933.9 (5.3) |  |
| First visit age, mean (SD) |  | 51.2 (11.4) | 57.0 (10.4) |  |
| Sex, n (%) | Female | 139548 (55.7) | 468 (62.5) |  |
|  | Male | 110829 (44.2) | 281 (37.5) |  |
|  | Nonbinary/Unknow |  |  |  |
|  | n | 168 (0.1) |  |  |
| R&E, n (%) | Asian/NHPI | 32427 (12.9) | 151 (20.2) |  |
|  | Black | 17111 (6.8) | 62 (8.3) |  |
|  | Latinx | 15036 (6.0) | 53 (7.1) |  |
|  | Other/Unknown | 28177 (11.2) | 45 (6.0) |  |
|  | White | 157794 (63.0) | 438 (58.5) |  |
| Matched Patients for -1 year model |  |  |  |  |
|  |  | Control | AD | SMD |
| n |  | 4184 | 523 |  |
| Birth year, mean (SD) |  | 1934.2 (5.6) | 1934.0 (5.3) | -0.042 |
| First visit age, mean (SD) |  | 57.2 (9.4) | 56.9 (10.5) | -0.028 |
| Prediction age, mean (SD) |  | 73.1 (5.8) | 73.1 (5.8) | -0.002 |
| Years in EHR, mean (SD) |  | 15.9 (7.8) | 15.9 (7.9) | -0.004 |
| Log(n prev visits), mean (SD) |  | 3.6 (1.5) | 3.7 (1.6) | 0.065 |
| Log(n concepts), mean (SD) |  | 3.1 (1.3) | 3.3 (1.4) | 0.108 |
| Log(days since first event), mean (SD) |  | 8.5 (0.4) | 8.5 (0.4) | 0.043 |
| Sex, n (%) | Female | 2343 (56.0) | 317 (60.6) | 0.094 |
|  | Male | 1841 (44.0) | 206 (39.4) |  |
| R&E, n (%) | Asian/NHPI | 705 (16.8) | 112 (21.4) | 0.219 |
|  | Black | 520 (12.4) | 35 (6.7) |  |
|  | Latinx | 280 (6.7) | 39 (7.5) |  |
|  | Other/Unknown | 223 (5.3) | 32 (6.1) |  |
|  | White | 2456 (58.7) | 305 (58.3) |  |

**Figure 1:**
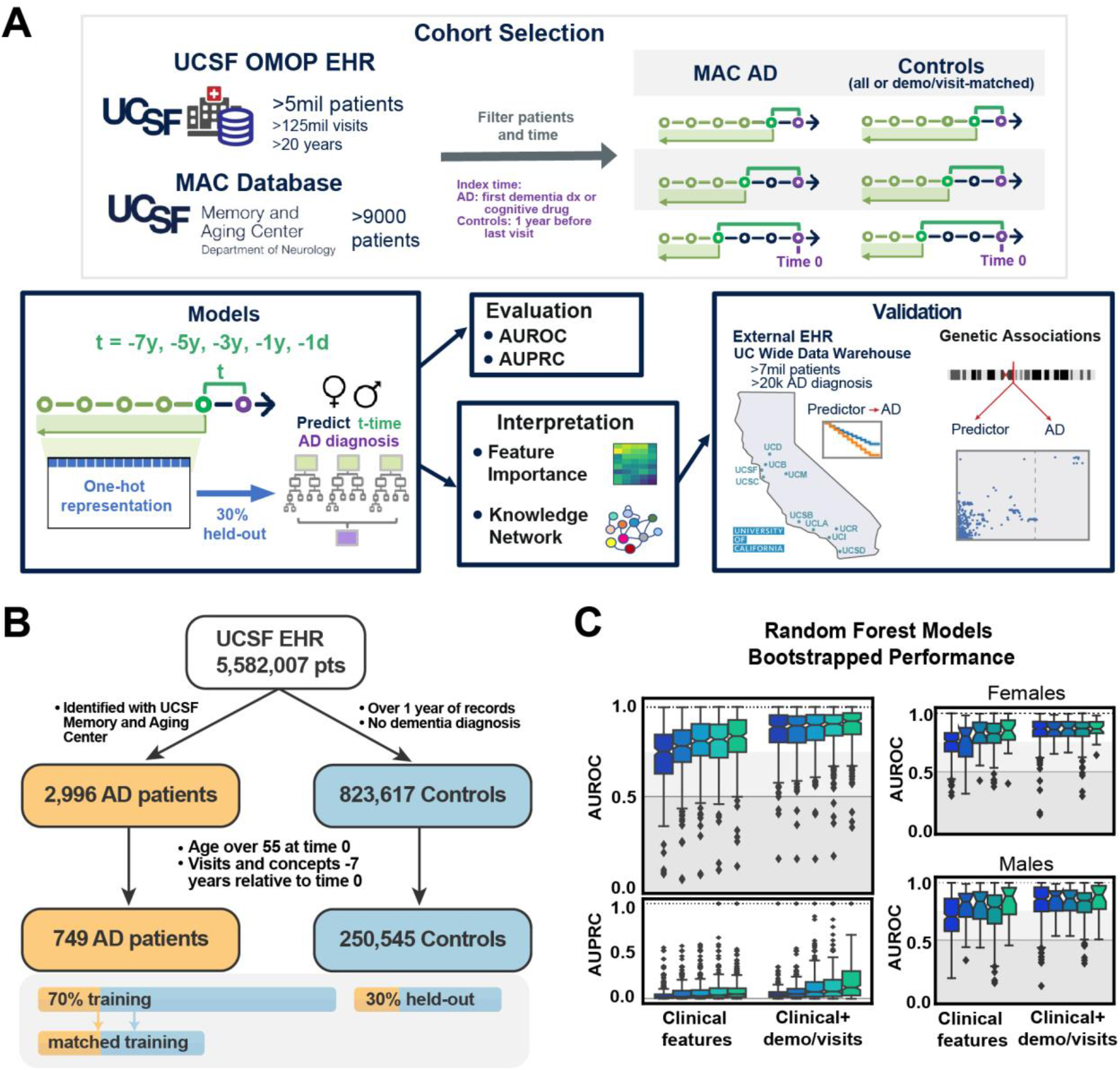
Overview of Patient Selection and Random Forest Model Performance. A. From the UCSF electronic health records and the UCSF Memory and Aging center database, patients and clinical information was extracted, filtered, and prepared for time points before the index time. All clinical features extracted were one hot encoded and trained on random forest models to predict future risk of Alzheimer’s Disease diagnosis. Models were evaluated on a 30% held-out test set to compute AUROC/AUPRC, and interpreted based on feature importances and using a heterogeneous knowledge network (SPOKE). Top features were then further validated in external databases. B. Filtering of a consistent set of AD and Control patients from the UCSF EHR for model training and testing. Filtered patient cohorts are shown in Table 1, and split with 30% held-out set for testing. C. Bootstrapped performance of random forest models on the full held-out test set (prevalence of AD on held-out set = 0.003). Bootstrapped AUROC performance for models trained and tested on female strata and male strata are also shown.

### ML models based on clinical data can accurately predict Alzheimer’s Disease onset up to 7 years in advance

Random forest (RF) models trained on only clinical features from time points between −7 years to −1 day to AD onset were evaluated on the held-out dataset with average bootstrapped Area Under the Receiver Operating Characteristic (AUROC) curve between 0.72 (median 0.75) for the −7 year time model to 0.81 (median 0.85) for the −1 day model. The RF models performed with Area Under the Precision Recall Curve (AUPRC) greater than the reference held-out test set AD prevalence of 0.003 (average/median of 0.05/0.01 for −7 year model and 0.10/0.06 for −1 day model, Figure 1C). With addition of demographics and visit-related features, RF model performance improved with average bootstrapped AUROC between 0.86 (median 0.89) to 0.90 (median 0.94) and AUPRC between mean 0.06 (median 0.04) and 0.27 (median 0.14) for the −7 year to −1 day model, respectively (Figure 1C).

Top features across each time point model (see Methods) included features across clinical data domains, including vaccines, abnormal feces content, hypertension, hyperlipidemia (HLD), and cataracts (Supplemental Figure 1A). Demographic and visit-related features became predictive for AD diagnosis when added to the model, which is not unexpected since these features may contribute to confounding that influence the identified features and predicted risk of AD diagnosis (Supplemental Figure 1A). EHR diagnoses mapped to phecode categories^29^ (see Methods) identified sense organs, circulatory, and musculoskeletal phecode categories for early models, and mental disorder category for late models (Supplemental Figure 1B). Among the clusters of top 50 ranked phecodes, one cluster identified phecode features that maintain high relative importance throughout the time models (HLD, hypertension, dizziness, abnormal stool contents), and other clusters contain features with relative importance at specific time points (Supplemental Figure 1C). While some of these features support prior identified AD risk factors, the lack of adjustment may lead to feature identification as proxies for age in risk determination but not directly relevant to disease pathogenesis. Therefore, we proceed to identify disease relevant features by training models on patients matched on demographics and hospital utilization for the goal of hypothesis generation.

### Models trained on matched cohorts can identify hypotheses for biologically relevant AD predictors

To train models that are robust for AD prediction for identifying predictors without demographic and visit-related confounding, we train time point models on a matched set of participants at a 1:8 ratio between AD and controls. Sufficient balance was achieved on numerical covariates that were highly important in unmatched demographic models (Supplemental Figure 2, Supplemental Table 3).

RF models trained on only clinical features from −7 years to −1 day performed with average bootstrapped held-out test set AUROC between .58 (median 0.57) for the −7 year time model to .77 (median 0.77) for the −1 day time model. The models performed with AUPRC greater than the held-out test set AD prevalence of 0.003 with improvement closer to time 0 (mean/median of 0.02/0.008 for −7 year time model and 0.08/0.03 for −1 day model, Figure 2A). When demographics and visit-related information were added as features, the models performed with minimal improvement, with average bootstrapped test set AUROC between 0.61 (median 0.61) to 0.71 (median 0.72) and similar AUPRC (mean/median of 0.02/0.009 for −7 year time model and 0.05/0.03 for −1 day model, Figure 2A).

**Figure 2:**
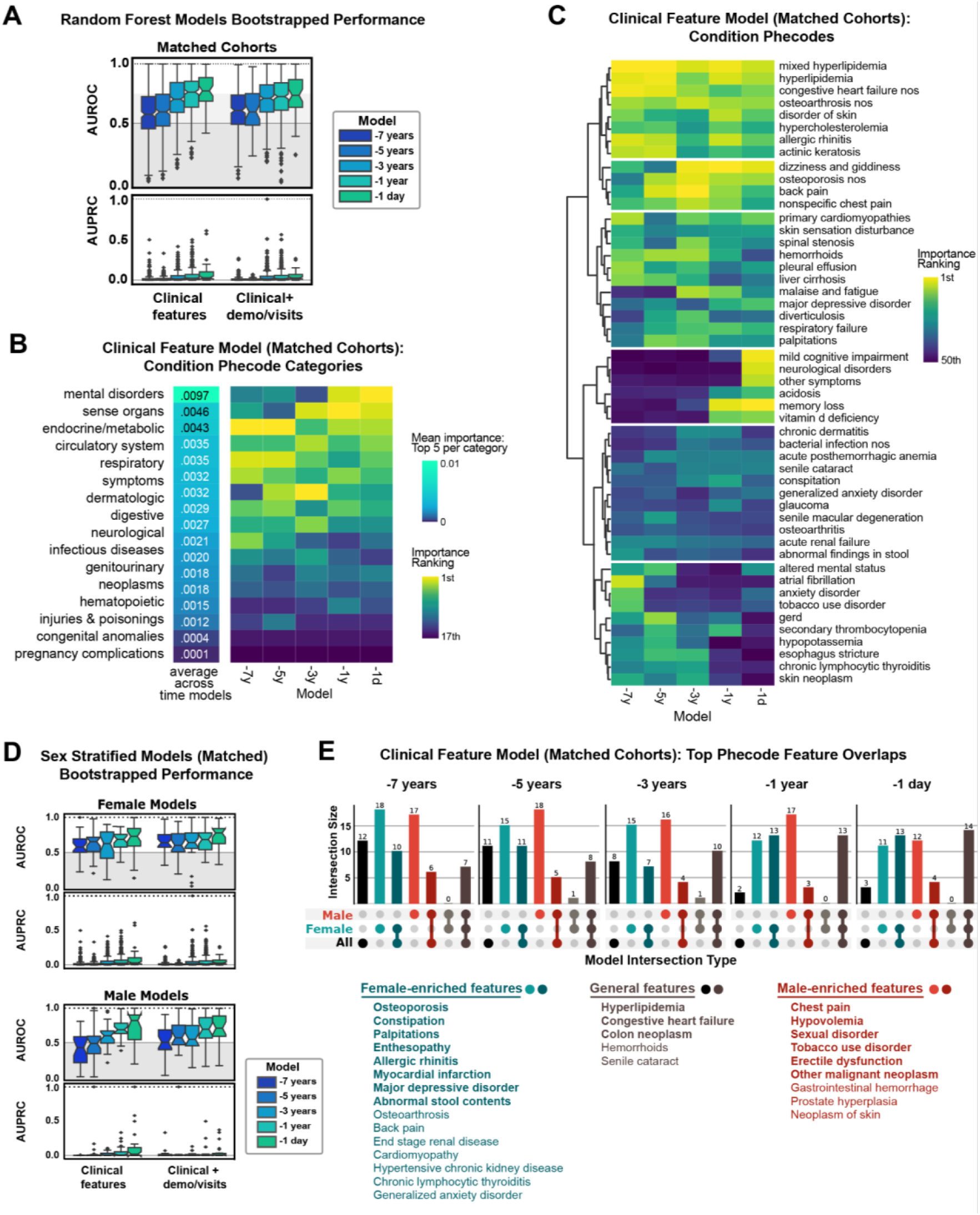
Models trained on matched cohorts allows for identification of hypotheses for AD predictors A. Bootstrapped performance of models trained on cohorts matched by demographics and visit-related factors on the full held-out test set (prevalence of AD on held-out set = 0.003). B. Top clinical phecode categories for matched models ranked by the average of the top 5 importance for each phecode category. Sorting is based on this average across time models. C. Top 50 phecodes (detailed features) across time models, with features clustered based on ward distance of rankings. D. Bootstrapped performances of sex-stratified matched models on the held-out test set (reference AUPRC = .0036 female, .0022 male). E. Overlap of top matched model features for models trained on all patients, female stratified patients, and male stratified patients, with model cutoff importance (RF average impurity decrease) greater than 1E-6. Specific features are listed, with bold features indicating top features across all 5 time models, and non-bolded features indicating top features across 4 time models.

Among top features sorted by average importance across time models, top features include amnesia and cognitive concerns, HLD, dizziness, cataract, congestive heart failure, osteoarthritis, and others (Figure 2B). These top features are consistently important even when demographics and visit information was added to the model, although demographic and visit features still had minimal influence on prediction (Figure 2B).

Since matching allows for the control of the influence of visit and demographic-related variables utilized in matching on AD prediction, the remaining diagnoses features can be identified for hypothesis generation with greater specificity for AD predictive risk. Top phecode categories include mental disorders, sense organs, and endocrine/metabolic categories (Figure 2C). Among clusters of specific phecodes, one cluster included features with maintained predictive importance throughout time models (HLD and congestive heart failure), while other clusters include phecodes that are relatively predictive several years prior to AD onset (osteoarthritis, allergic rhinitis). A cluster of features emerges as important around −3 years (osteoporosis, dizziness, back pain, hemorrhoids, palpitations), and some features only emerge as important closer to the time of AD onset (memory loss, vitamin D deficiency, Figure 2C). Together, this shows that the model can identify a combination of conditions that can lead to AD risk identification for a patient of a given age and hospital utilization burden.

### Stratification by sex allows identification of features that are predictive within a subgroup

Since sex plays a role in AD risk, models were trained within male or female-identified sex groups to perform sex-specific prediction and identify sex-specific predictive features, without and with matching on demographics and hospital utilization (demographics in Supplemental Table 4). Models trained on clinical features performed with average held-out test set AUROC between 0.75 (median 0.76) and 0.71 (median 0.71) for −7 year female and male models to 0.84 (median 0.86) and 0.82 (0.89) for −1 day female and male models. For AUPRC, the models performed greater than the held-out test set prevalence (0.0036 for females, 0.0023 for males) with performance of 0.056-0.11(median 0.022-0.061) and 0.041-0.15(median 0.015-0.056) for females and male −7 years to −1 day time models respectively. With addition of demographics and visit-related features, AUROC/AUPRC improved considerably (Supplemental Figure 3A).Top features include sense organs and musculoskeletal phecode categories in female-only models, and circulatory system and digestive phecode categories as important among male-only models (Supplemental Figure 3B).

To identify sex-specific biologically relevant clinical predictors for hypothesis generation, models were also trained by matching on demographic and visit-related factors within each subgroup (matching results in Supplemental Table 4). Time point models trained only on clinical features performed with mean held-out test set AUROC between 0.60-0.68 (median 0.58-0.74) and 0.41-0.75 (median 0.43-0.84) for female and male models respectively (Figure 2D). For AUPRC, models performed greater than held-out test set prevalence with performance ranging from 0.031-0.095 (median 0.0076-0.046) and 0.0040-0.125 (0.0033-0.022) for female and male models respectively. Slight improvement in performance was observed with the addition of demographics and visit-related information (Figure 2D).

Top phecode categories in the female models include respiratory/circulatory system features earlier on, to musculoskeletal features in the −5 year model, to sense organs and mental disorders in the −1 year and −1 day model. Top categories in male models include endocrine/metabolic/circulatory disorders earlier, to digestive and genitourinary in −5 and −3 models, to mental disorders in −1 day model (Supplemental Figure 3B). When comparing specific phecodes, some are general across the subgroups such as HLD, congestive heart failure (early models), and memory/cognitive symptoms (later models) (Figure 2E, Supplemental Figure 3C). Female-driven features across time models included osteoporosis, palpitations, allergic rhinitis, myocardial infarction, major depressive disorder, and abnormal stool contents. Male-driven features included chest pain, hypovolemia, sexual disorder, tobacco use disorder, and neoplasms (Figure 2E).

### Use of a knowledge graph allowed identification of potential biological explanations underlying predictive features

Next, we utilized the SPOKE knowledge graph^28^ in order to explain potential biological relationships between top clinical model features and AD. We mapped biological features (genes, proteins, compounds, etc.) between top 25 clinical predictors (mapped to disease nodes) and AD node for each model (see Methods).

Genes that appear in shortest path networks among matched models across multiple time include APOE, AKT1, INS, ALB, IL1B, INF, ALB, IL6, SOD1, etc. and compounds include atorvastatin, simvastatin, ergocalciferol, progesterone, estrogen, cyanocobalamin, and folic acid (Figure 3). These genes and compounds also share relationships to multiple occurring model input nodes, particularly familial hyperlipidemia and osteoporosis among all time point models (Figure 3). Notable nodes that appear over at least 2 models include C9orf72, TREM2, APP, MAPT with relationships to input nodes of musculoskeletal and joint disorders, deafness, and depression (Figure 3).

**Figure 3:**
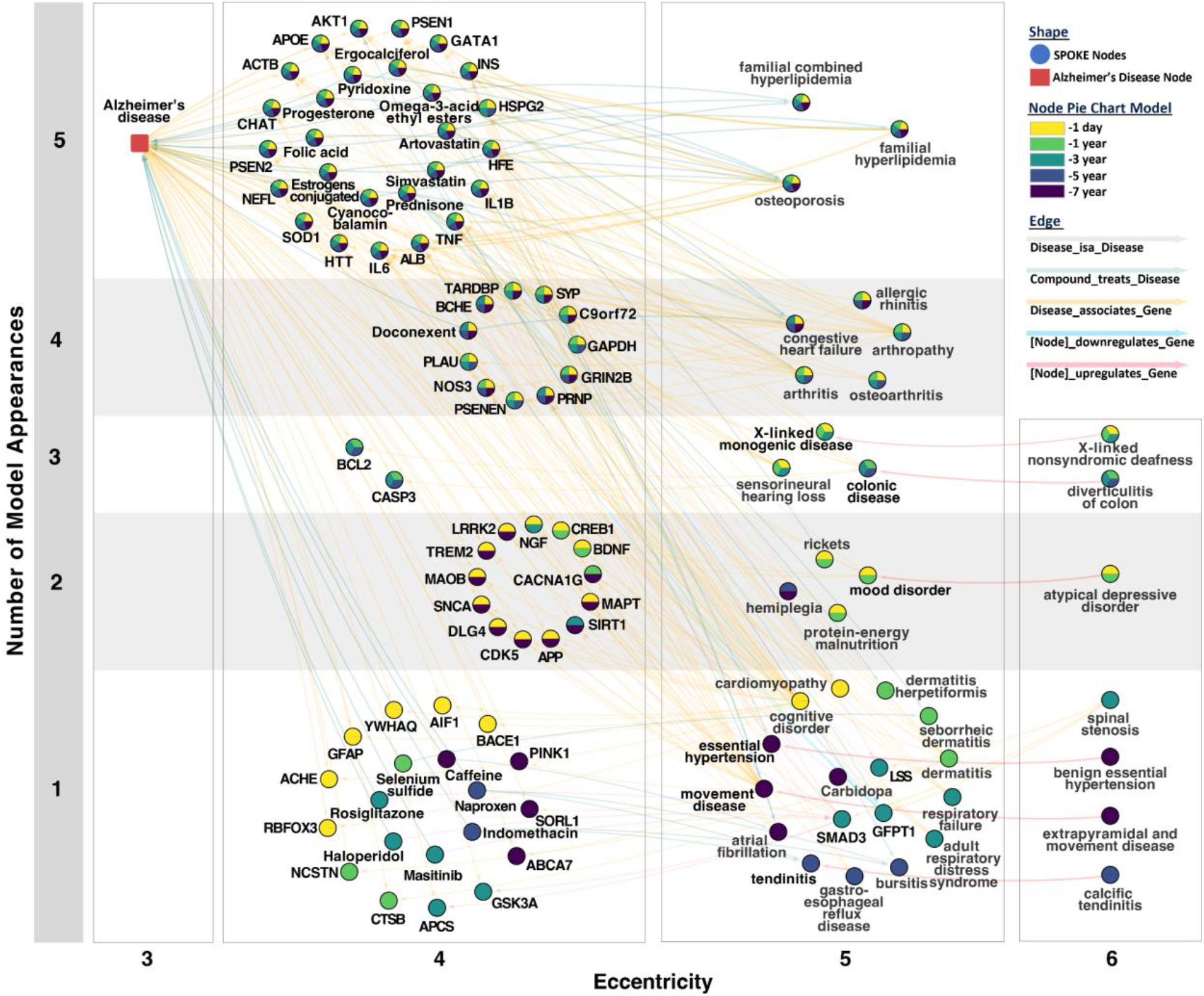
SPOKE provides biological interpretation of hypotheses associated with shared clinical phenotypes Combined SPOKE network of all shortest paths to Alzheimer’s Disease node (DOID:10652) for top 25 input features from matched AD model at every time point. Network is organized based on the number of time point occurrences (y-axis) and eccentricity of a node in the subnetwork (x-axis). Specific time point occurrences are colored by the pie chart within each node.

### Hyperlipidemia validates as a top predictor of AD in external EHRs and a genetic link confirmed in APOE locus

In order to further validate the utility of models to identify predictive disease associations, we followed up on HLD as a top feature that was a consistent predictor across all models. Utilizing a retrospective cohort study design in an EHR on five hospitals across the University of California system (University of California Data Discovery Platform (UCDDP)) with exclusion of UCSF, HLD-diagnosed patients (exposed group, n = 364,289) had a faster progression to AD event compared to matched unexposed patients (n = 364,289, matched demographics in Supplemental Table 6) (Figure 4A, Supplemental Figure 4A, log-rank test p-value<0.005). This was further confirmed with a Cox proportional hazards analysis (hazard ratio (HR) 1.52 (95% Confidence Interval (CI) 1.46-1.57), visit/demographic adjusted HR (aHR) 1.26 (1.21-1.31), p-value <0.005, Supplemental Figure 4C).

**Figure 4:**
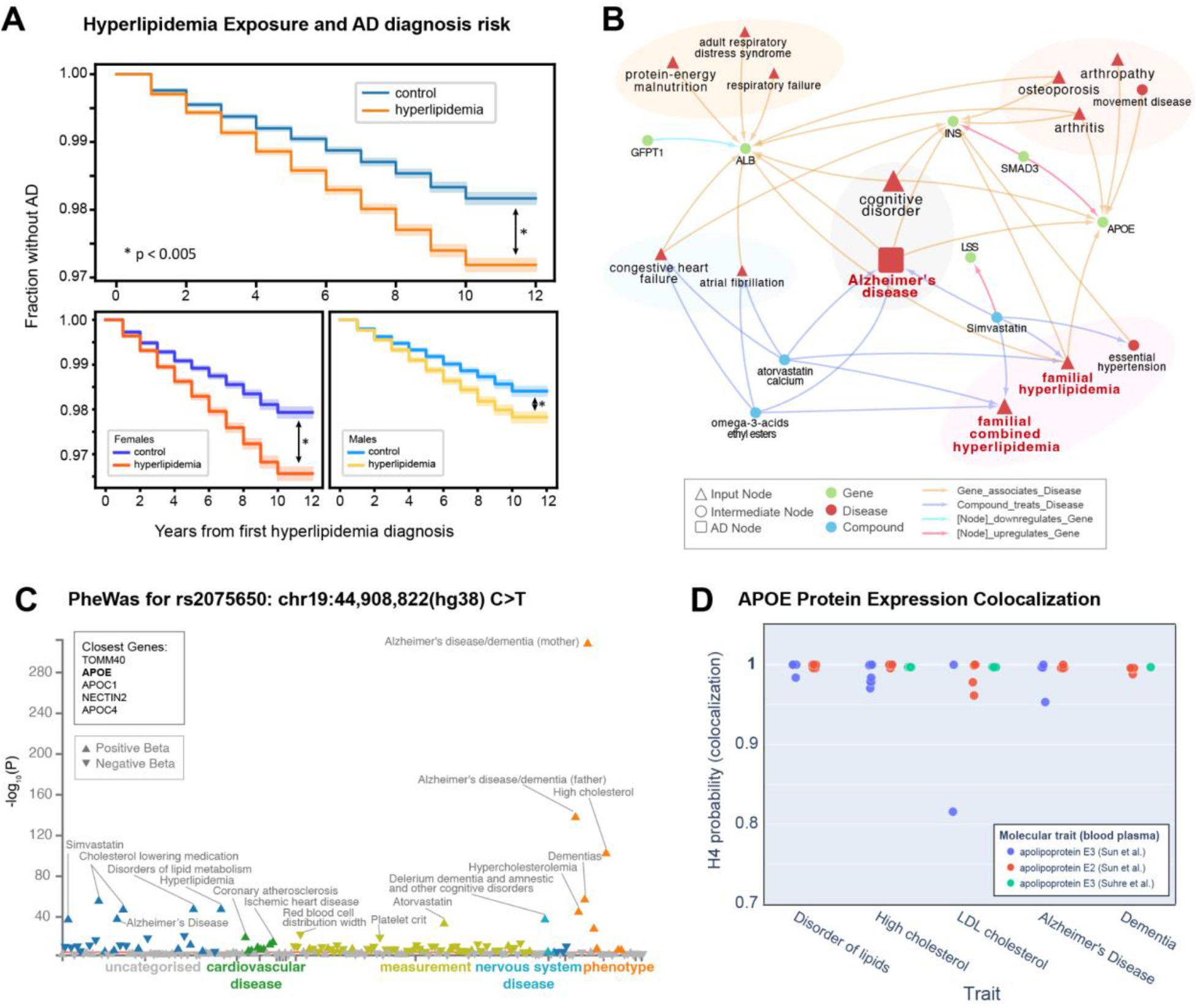
The hyperlipidemia and AD association is validated externally with APOE as a shared causal genetic link A. Kaplan Meyer curve on UC-wide EHR for hyperlipidemia (HLD) as the exposure. Log rank test is significant for all HLD vs controls (p=2.36e-85), female HLD vs female controls (p=3.64e-69), and male HLD vs male controls (p=8.39e-22). B. 1st and 2nd degree neighbor of hyperlipidemia on the full network representing all shortest paths from the top 25 features per time model. C. PheWAS for variant rs2075650 on a shared loci associated with both hyperlipidemia and AD, plotted based on associations with phenotypes in the UK Biobank. D. Plot of APOE protein expression colocalization with H4 (probability two associated traits share a causal variant) from Open Targets Genetics. Each dot represents a specific phenotype categorized based on trait (x-axis). Each color represents an APOE molecular trait measured from blood plasma from Sun et al. and Suhre et al.

In order to investigate potential relationships between HLD and AD, the HLD-specific knowledge network demonstrated shared gene associations with LSS, APOE, INS, SMAD3, ALB, and GFPT1 (Figure 4B). Locus intersections between high LDL cholesterol and AD across two independent GWAS studies across 408,942 AD patients from Schwartzentruber et al.^30^ and 94,595 LDL Cholesterol patients from Willer et al.^31^ respectively identified multiple shared variants, including ch19:44,892,362(hg38):A>G (rs2075650) and ch19:44,905,579(hg38):T>G (rs405509)(genetics.opentargets.org/study-comparison/GCST002222?studyIds=GCST90012878). PheWAS for rs2075650 on the UK Biobank verified significant associations with cholesterol levels, HLD, AD, and family history of AD (Figure 4C). Colocalization H4 probability, a measure that determines the probability two traits are associated at a locus based on prior genetic studies, supports a causal link with locus variants for APOE protein QTL and both HLD traits and AD traits (Figure 4D).

### Female-specific predictor of osteoporosis validates in an external EHR with potential explanations given in SPOKE and genetic colocalization analysis

Osteoporosis was identified as an important feature in the matched models as a female-specific clinical predictor of AD. In the UCDDP, osteoporosis-exposed patients (n=68,940) showed a quicker progression to AD compared to matched unexposed patients (n=68,940, matched demographics in Supplemental Table 7) (Figure 5A, Supplemental Figure 4B, log-rank test p-value<0.005). When stratified by sex, this progression is significant when comparing between female osteoporosis (n=57,486) vs female controls (n=58,636). Cox hazard analysis further supported osteoporosis as a general risk feature for AD (HR 1.81 (95% CI 1.70-1.92), aHR 1.59 (1.45-1.70), p<.005 Supplemental Figure 4D).

**Figure 5:**
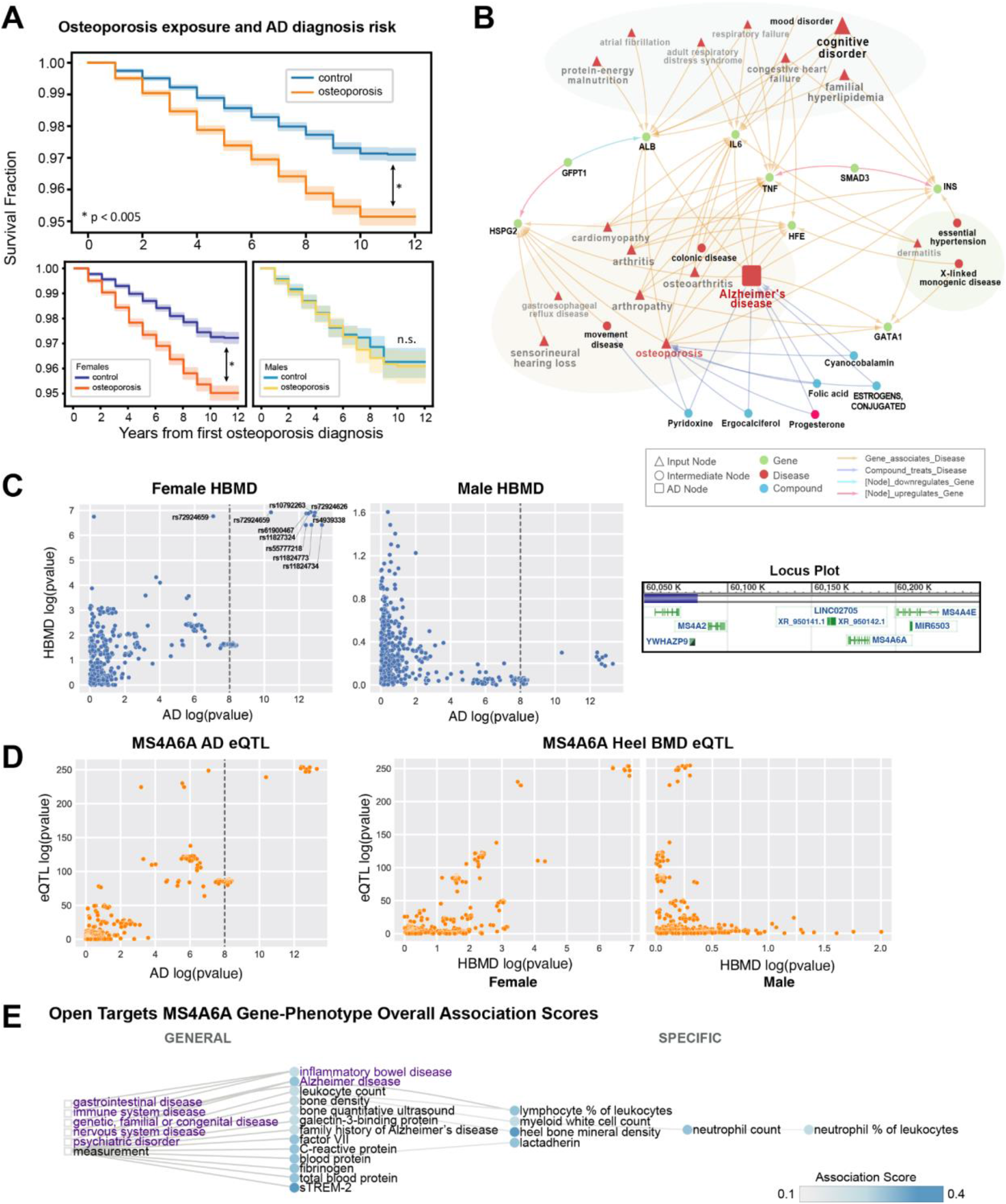
The association between osteoporosis and AD is validated externally with MS4A6A as a potential female-specific shared genetic link A. Kaplan Meyer curve on UC-wide EHR for osteoporosis as the exposure. Log rank test is significant for all osteoporosis vs controls and similarly for female strata, * p<0.005. B. 1st and 2nd degree neighbors of osteoporosis node on the network representing all shortest paths from top 25 feature per time model. C. P-P plots between Alzheimer’s Disease GWAS (Jensen et al. 2018, n= 455,258) and sex-stratified heel bone mineral density GWAS (Female n = 111,152, Male HBMD n = 166,988, UK Biobank / Neale’s Lab GWAS) around the MS4A locus (left and middle plots) at region 60050000-60200000 of Chromosome 11 (locus plot on right). D. MS4A6A cis eQTL association with AD, and association with sex-stratified heel bone mineral density, from eQTLGen. E. Open Targets associated phenotype graph for MS4A6A with association score computed based on a weighted harmonic sum across evidence (described in platform-docs.opentargets.org/associations#association-scores). Purple words indicate diseases, while black words indicate measurements. Circles are phenotypes colored by the association score, and boxes represent the most general categories.

Osteoporosis-specific SPOKE network demonstrated shared gene associations with IL6, SMAD3, TNF, HSPG2, GATA1, GFPT1, HFE, INS, and ALB (Figure 5B). Based on previous GWAS studies across 472,868 AD patients from Schwartzentruber et al.^30^ and 426,824 heel bone mineral density (HBMD) patients from Morris et al.^32^, a shared risk locus was found in Chromosome 11 between HBMD and AD among the MS4A gene family (https://genetics.opentargets.org/study-comparison/GCST006979?studyIds=GCST90012877), with the closest gene as MS4A6A. A comparison of prior GWAS of up to 71,880 AD patients from Jansen et al.^33^ and sex-stratified heel bone mineral density (HBMD) GWAS (111,152 Female, 166,988 Male) of UK Biobank patients from Neale Labs (www.nealelab.is/uk-biobank/) supports a female-specific association at the shared locus (Figure 5C). Colocalization analysis supports a link between MS4A6A and AD (H4 = 0.987), female-specific HBMD with AD, and phenotypes with MS4A6A expression (Figure 5D, AD vs Female HBMD H4 = 0.998, MS4A6A vs Female HBMD H4 = 0.997). This statistical significance is not replicated for male specific HBMD GWAS (Figure 5D, AD vs Male HBMD H4 = 0.00263, MS4A6A vs Male HBMD H4 = 0.00266). MS4A6A weighted associations with other phenotypes from Open Targets Genetics found locus associations with many inflammatory phenotypes including c-reactive protein, lymphocyte percentage, and neutrophil count (Figure 5E).

## Discussion

While there is great potential in ML on clinical data, balancing clinical utility and biological interpretability can be challenging. Cohort selection and data preprocessing is a crucial first step. To address this, we used thousands of EHR concepts to develop prediction models for expert-identified AD diagnosis, and selected an index time suggesting AD onset. Our general prediction model shows predictive power up to −7 years before the defined index time of AD onset. This model can help assess disease risk before time-consuming and costly detailed neuropsychological, biomarker, or neuroimaging assessments. The model may also identify at-risk patients for follow-up or inclusion in early intervention or clinical trials. Furthermore, interpretable models allow clinicians to understand what clinical features were used in determining prediction probability and assess the model output with greater trust compared to “black box” models.

In order to identify early clinical predictors that may be biologically relevant for AD diagnosis, we trained models on patients matched by pre-identified confounding variables such as demographics and visit-related features so that these features have less influence in AD prediction. Machine learning models still retain the ability to predict AD diagnosis with mean AUROC over .70 after the −3 year time model for random forests. Inclusion of demographic and visit-related features minimally improved model performance, which is expected since matching increased the specificity of the task to predict AD onset controlled on demographics and visit-related features. In terms of clinical utility, the models trained on matched patients provide predictive power for a given clinical scenario between two patients with similar pre-test probability of AD risk (e.g. same age and disease burden), with application of this model as a tool for determining post-test probability of future AD risk. Furthermore, by balancing on pre-identified confounders such as demographics and visits, top features may be interpreted with more biological relevance for AD risk. For example, while we identified essential hypertension as an important feature in the models trained on the full cohort, this diagnosis became less important in the models trained on matched cohorts, suggesting hypertension may be nonspecific for AD and may instead be more directly related to aging or disease burden.

Our time models trained on matched cohorts identify or strengthen known or suggested hypotheses for early clinical predictors of AD, such as hyperlipidemia as a feature for all time point models. We also identify relative importance of features years in advance, such as allergic rhinitis and atrial fibrillation as early predictors, osteoporosis and dizziness as non-neurological predictors, and cognitive impairment and vitamin D deficiency as late predictors of AD. These findings potentially support hypotheses suggesting AD can be associated with general aging or frailty, which might present in non-neurologic body systems either prior to or concurrent with AD ^34–38^. Nevertheless, while these models can identify hypotheses of predictive features, EHR data can still capture clinical biases or misdiagnoses, and further studies can investigate the influence of behavioral bias vs biological relevance.

We further trained models on sex-stratified subgroups (female vs male), with and without matching on demographics and visit-related covariates, in order to identify sex-specific drivers of clinical predictors. Given evidence that sex may influence different pathways to AD diagnosis^22,39,40^, it is important to consider how patient heterogeneity may impact the training, utility, and interpretation of a prediction model. From the matched cohort models, we identified clinical features in each subgroup that were consistent with the general models, such as hyperlipidemia as important in every model and memory loss as important in late models. Furthermore, we identified features that were sex-specific, such as osteoporosis, depression, allergic rhinitis, and abnormal stool contents as predictors enriched among women, and chest pain, hypovolemia, and prostate hyperplasia as predictive among men. Further work can seek to disentangle the biological meaning of these sex-specific predictive features: whether they reflect sex-specific non-neurological manifestation of prodromal states, contributing risk factors, or even sex biases in clinician evaluation and treatment (e.g., bone density evaluation may arise more often after a fall). These models also demonstrate that for a heterogeneous disorder like AD, subgroup composition, like sex ratio of a cohort, can influence features that are identified as important. For example, the higher preponderance of females leading to sex-specific predictive factor, osteoporosis, being identified as a general predictive variable in the general group. This further indicates that both generalizable models and subgroup-specific models can provide valuable insight, both general and personalized, for a complex disease.

We investigated shortest paths between top model predictors and heterogeneous networks (SPOKE) in order to identify biological hypotheses that may explain the relationship between identified early clinical phenotypes and AD. This allowed us to identify multiple genes that may be relevant that can give insight into the high order constellation of clinical symptoms and AD. For example, we were able to identify known genetic associations with dementia based upon symptoms from the rest of the body, such as through identification of known autosomal dominant early AD genes such as APP and PSEN 1/2^41^. Other genes identified with possible associations with AD include APOE, HFE, and HSPG2 variants that impact AD risk^42–46^. While these associations are included in the SPOKE network due to evidence in literature, the association of these genes with other early clinical predictors is less established, and thus this analysis allowed us to identify a novel constellation of phenotypes observable in a clinical setting that can lead a clinician to suspect future AD risk.

To validate top clinical predictors, we utilized a hypothesis-driven approach to investigate the relationship between two identified features (hyperlipidemia and osteoporosis) and progress to AD diagnosis in an external database across the University of California EHR system. For both phenotypes, the UC-wide EHR database supports a potential increased AD diagnosis risk due to evidence of decreased time to AD and increased hazard of AD diagnosis in patients exposed to the predictor of interest. The association between hyperlipidemia and AD has been identified in prior clinical studies and systematic reviews^47–50^. In particular, APOE is a well-established associated genetic locus^51^, and APOE polymorphism is known to modify AD risk, particularly in individuals carrying the ε4 allele^52^. Many studies have also shown APOE association with elevated lipid levels and cardiovascular risk factors^53,54^. The validation of these well-known associations not only show that our ML models on clinical data can pick up hyperlipidemia as a risk factor, but also by utilizing the SPOKE network we can integrate known relationships in literature to potentially explain the association between hyperlipidemia and AD and identify the APOE locus as a potential shared causal mechanism as demonstrated in the colocalization results. Beyond the ability to identify known relationships, the SPOKE network also proposes biological explanations of higher-order shared associations between clinical predictors, such as ALB as a shared genetic association between congestive heart failure, malnutrition, hyperlipidemia, and AD, or INS as a shared association between osteoporosis, hypertension, hyperlipidemia, and AD. Prior studies have identified potential mechanisms underlying the relationship between energy utilization, lipid levels, nutrition, and neurodegeneration^55–57^, although specific hypotheses of mechanistic relationships are an area for exploration in future studies.

The association between osteoporosis and AD is also validated to a lesser extent in clinical studies and meta-analysis^58,59^, with unclear but possible sex-modification of this effect. Our study identifies osteoporosis as a predictor for AD among females prior to AD, but shows less of a *relative* predictive effect for males compared to other clinical features. Nevertheless, it is still possible that shared relationships between osteoporosis and AD exist in males. A bone mineral density GWAS analysis of female patients shows p-value association with AD GWAS around the MS4A family locus, and this is further supported by MS4A6A eQTL colocalization with both Alzheimer and female HBMD. Prior studies have established the MS4A gene cluster as a risk for AD, with one study identifying the cluster based on mendelian randomization^60^, and another that identifies a stronger female-specific effect size for MS4A6A^61^. Some studies investigating the role of the MS4A family suggest mechanisms that involve immune function, particularly among microglia^62^. While this gene may not have been identified in SPOKE, SPOKE did capture direct pathways through known markers of inflammation such as IL6 and TNF, and we also see MS4A6A as highly associated with measurements of immune cells in the blood. Further studies will be needed to validate the exact associative mechanism between osteoporosis and AD, although some prior hypotheses suggest the potential impact of genetic variants on osteoclast function, amyloid clearance, or oxidative stress response^63,64^.

This study has several limitations. First, EHR data complexity and quality can affect prediction models, and it is challenging to distinguish the influence of clinician/patient behavior, sociological factors, or underlying biology on identification of features. Matching can improve interpretability by removing influence of non-biological covariates, but follow-up validation of hypotheses across omics data types is needed. Due to changing patient demographics and societal factors, prediction models should be continuously trained, updated, and evaluated if implemented in the clinical setting to ensure effective utilization and account for biases that may have been learned from the data. Second, clinical EHR data is sometimes sparse and provides a superficial interval snapshot of a patient’s health, so the absence of a record may not necessarily reflect the absence of a condition. Third, survival models have extensive right censorship and do not take into account competing risks. Fourth, since AD is heterogeneous and differential diagnosis is nuanced and subjective even in expert hands, predictive performance can be limited by label quality and the signal from clinical features can be noisy, limiting performance and generalizability. Future work investigating heterogeneity may identify subgroup-specific features where subgroups can be divided based on biotype, dementia syndromes, racialization, and so on. Future applications with hierarchical models, transfer learning, or fine-tuning on a subpopulation can increase personalization of models. Fifth, our sex-stratified analysis was restricted to patients that identified as female or male. Future studies could explore AD patterns among intersex individuals. Lastly, predictive features identified are relevant prior to AD onset, and future work is needed to identify diagnostic-relevant AD comorbidities, or conditions that can occur after AD progression. Since predictive features are identified as hypotheses, the direct mechanism and causal pathway relating a phenotype to AD is not known. Future work can investigate causality with mendelian randomization or mechanistic studies.

In this study, we demonstrate how formulation of prediction models can influence utility for predictive application or biological interpretation. We show how models can be utilized to identify early predictors, and utilize SPOKE to explain relationships via shared biological associations. Lastly, we show that our models can pick up known associations with HLD through APOE, and identify a lesser known association with osteoporosis through MS4A6A that may be female-specific. This study contributes to the field of EHR integrative research that can inform future directions in both AD care and research.

## Methods

### Patient Identification

Alzheimer’s Disease (AD) patients were identified based on UCSF Memory and Aging Center database containing over 9000 patients mapped to the UCSF OMOP-format EHR. These patients have undergone dementia evaluation at the Memory and Aging Center and thus had expert-level clinical diagnoses. The remaining control patients were obtained from the rest of the UCSF EHR, with over 1 year of records and no existing records of dementia diagnosis among the G[123]* ICD-10 categories (Supplemental Table 1).

An index time was determined to filter the input features prior to first clinical indication of dementia. This was defined among the AD cohort as the first time of any AD diagnosis, dementia diagnosis, or prescription of cognitive drug (ATC codes N06D, Supplemental Table 2) as determined to be the first time point of AD manifestation. For controls, the index time was defined as 1 year before the last recorded EHR visit date. In order to maintain a consistent patient population for training and evaluation of machine learning models, the final AD and control cohort was identified by filtering to patients who are at least 55 years of age at the index time and have existing clinical visits and concepts 7 years prior to the index time.

### Data Extraction and Preparation

Demographics (birth year, gender, race, ethnicity), clinical concepts (conditions, drug exposures, abnormal measures), and as visit-related features (age at prediction, first visit age, years in UCSF EHR) were extracted before the index time for the AD and Control cohort from the UCSF Observational Medical Outcomes Partnership (OMOP) EHR database. Abnormal measures were extracted from the OMOP *measurement* table based on the numeric value falling either above *range_high* or below *range_low*.

To train models in advance of the index time, clinical information was extracted for each patient including all data up to a time point X before the index time, where X includes −7 years, −5 years, −3 years, −1 years, and −1 day. These time points represent the knowledge of a patient’s clinical history leading up to time X before time 0, which represents the knowledge of a clinician when a patient comes for a visit. All existing clinical features (conditions, drugs, measurements) were one-hot encoded. Demographic and visit-related features (prediction age, first visit age, years in UCSF EHR, log(number prior visits), log(number prior concepts), log(days since first clinical event)) were scaled between 0-1 on the training data (log indicates natural logarithm, feature scaling allows for multiple ML model approaches). All features with no variance were removed.

### Machine Learning Preparation and Training

Random forest (RF) binary classification time point models for AD were trained using the patient representation at each time point before the index time. Training was performed on the 70% split, and the 30% held-out test set was utilized to evaluate the performance of all models. Models were trained with clinical features only (clinical model) and with clinical features + demographics and visit-related information (clinical + demo/visits model). For the models trained on matched samples, control patients were matched to AD patients at a 1:8 ratio on demographics and visit-related information utilizing propensity score matching^65^ (propensity score estimated based upon a logistic regression model, nearest neighbor matching without replacement).

Random forests were trained using *scikit-learn* package^66^, with balanced class weight parameter. Hyper-parameters were tuned (grid search) based on cross-validation performance (5 folds) of AUROC to determine parameters of *n_estimators* (n_features, n_features*2, n_features*3), *max_depth* (3, 5, None), and *max_features* (sqrt, log2). The number of estimators and max depth were tuned to balance between performance and overfitting, while a subset of features (max_features) was utilized per tree to help account for high correlation between features^67,68^. Models were evaluated on bootstrapped subsamples (50-200 iterations, 1000 samples) of the 30% held-out test set and evaluated on AUROC (area under the receiver operating curve) and AUPRC (aura under the precision-recall curve).

#### Stratification

Both models for full patient cohorts and matched cohorts were re-performed in sex strata in the same fashion (female strata, male strata). AD patients were re-matched to controls within each strata for the matched patient trained models. Models were evaluated similarly based on AUROC/AUPRC on the bootstrapped held-out test set.

### Top Feature Interpretation

Random forest models were investigated for feature interpretation due to the combined interpretable nature of the models (compared to neural networks) and the ability to capture nonlinear relationships (compared to logistic regression models)^69^. Average gini impurity decrease for each feature was utilized to evaluate the importance of each feature in the random forest models (feature importance). The average importance for each feature was taken across each time point models (-7yr, −5yr, −3yr, −1yr, −1day) to obtain an across-model importance for each model type, and normalized by the maximum importance value across all time point models within each model type (e.g. random forest) and group (e.g. female strata). Feature importances are then ranked within each model to obtain relative importance within each of the time points.

Since a patient’s exposure to a medication or a laboratory test is often a result of a diagnosis, we pursued interpretability based on diagnostic features that have been mapped to phecodes, which is a semi-manual hierarchical aggregation of meaningful EHR phenotypes^29^. This allows for a lossy categorization of detailed OMOP features (OMOP IDs) to phecodes (OMOP ID → SNOMED → ICD10 → phecode) and phecode category. SNOMED IDs were mapped to ICD10 based upon recommended rule-based mappings from the National Library of Medicine (NLM) September 2022 release (www.nlm.nih.gov/healthit/snomedct/us_edition.html). ICD10 codes were then mapped to phecodes based on the release from Wu et al.^70^ To obtain the importance within each phecode or phecode category, the average importance for the top 5 detailed OMOP features per phecode or phecode category was computed, and ranked between phecodes or categories. For phecodes across all models and sex-stratified models, the ranking of importance of phecodes across each time model was hierarchically clustered with Ward linkage.

To compare top phecodes between sex-stratified models to identify sex-specific features, top random forest features over an average importance threshold of 1e-6 were identified per time model trained on matched participants. Upset plots were then generated for each time point based upon this overlap. Female-driven features are defined as features that exist in both the full model and female models, or only female models, and male-driven features defined analogously.

### UC-wide validation analysis with hypothesis-driven retrospective cohort analysis

Two top clinical features were selected from the matched all patient model (hyperlipidemia) and matched sex-specific models (osteoporosis) and further followed up on an external EHR database to validate the feature as predictive and conferring risk for AD diagnosis. With these features defined as exposures, hypothesis-driven analysis was performed with a retrospective cohort study design on the University of California hospital EHR database (University of California Data Discovery Platform (UCDDP)) with exclusion of any patients seen at UCSF, so with included institutions consisting of UC Davis, UC Los Angeles, UC Riverside, UC San Diego, and UC Irvine. Exposed patients were identified with the exposure (hyperlipidemia or osteoporosis) and with recruitment age as the age of exposure diagnosis. Exposures were identified by string-matching and mapping to all descendants or related concepts based on the OMOP relationship tables, and final SNOMED codes are shown in Supplementary Information Controls were identified among the remaining patients and with recruitment age defined as the first visit age in the visit_occurrence table. All patients are then filtered to have at least 2 years of records in the EHR, and last visit age was utilized for right censorship.

The outcome of interest was AD diagnosis, which was identified based on SNOMED codes 26929004, 416780008, 416975007 (Supplementary Information 7). Exposed and control (unexposed) groups were then matched based on demographics (gender, race, ethnicity), birth year, and recruitment age. Analysis of time to AD diagnosis includes Kaplen Meyer survival analysis with log-rank test to compare survival curves between groups, and cox proportional hazard models were utilized to obtain unadjusted hazard ratios (HR) and adjusted hazard ratios (aHR) by demographics, visit information, with and without stratification by recruitment age or birth year.

### Heterogeneous Network analysis

Heterogeneous knowledge networks, such as SPOKE, integrate known relationships across biological and phenotypic data realms in databases and literature. Such a network could provide hypotheses to explain relationships between phenotypes that may not be immediately known^21,26^. We proceed with interpretation on the matched models, with the top 25 model features taken per time point and mapped to SPOKE nodes based on Nelson et al.^27^ Note that mappings may not be 1 to 1. All shortest paths were then computed from each input node to the Alzheimer’s Disease node (DOID: 10652), and shortest paths were filtered to exclude certain node types (Anatomy, SideEffect, AnatomyCellType,Nutrient) and edges (CONTRAINDICATES_CcD, CAUSES_CcSE, LOCALIZES_DlA, ISA_AiA, PARTOF_ApA, RESEMBLES_DrD). Edges were also filtered based on the following criteria: TREATS_CtD at least phase 3 clinical trial, UPREGULATES_KGuG/DOWNREGULATES_KGdG p-value at most 1E-4, PRESENTS DpS enrichment at least 5 and fisher p-value at most 1E-4.

If multiple detailed OMOP features map to the same node, the importance of the node was obtained by the average of OMOP feature importances. Networks for all time models were combined into a single network (union of nodes and edges), and total node importance was determined by the maximum across time. Network metrics were then computed with Cytoscape ‘Network Analyzer’ function^71^. The combined time model networks were then sorted by eccentricity metric on the x-axis (representing maximum distance to all other nodes, with lower number representing higher importance) and number of individual time model network occurrences in the y-axis (showing node importance persistence across time). Note that due to heterogeneous nature of edges and lack of edge weighting, distance in the figure is not meaningful.

To focus on two selected features for the full matched model (hyperlipidemia (HLD)) and the female-specific matched model (osteoporosis), the combined network was filtered based on first and second degree neighbors of the starting feature of interest. This allows for visualization of associated genes and AD, as well as relationships with other top model features found from the clinical models.

### Validation with Genetic Datasets

We further explored the association between clinical predictors and AD by identifying shared genetic loci between top model phenotypes and AD, based on colocalization probability and weighted evidence association scores computed from Open Targets Genetics^72,73^ (genetics.opentargets.org). Colocalization analysis is a method that determines if two independent signals at a locus share a causal variant, which helps increase the evidence that the two traits (e.g. hyperlipidemia and AD, or protein expression and AD) also share a causal mechanism. It is a Bayesian method which, for two traits, integrates evidence over all variants at a single locus to evaluate the following hypothesis that two associated traits share a causal variant. This is the H4 probability.

We first identified shared loci between the selected phenotypes (HLD or osteoporosis) and AD by identifying the genetic intersection between AD and related phenotypes in Open Targets Genetics.

For HLD and AD, we utilized the Open Targets Genetics platform to identify overlapping variants and shared locus between LDL Cholesterol and Family History of AD or AD. PheWAS between a shared SNP and UK Biobank phenotypes were plotted and extracted from the Open Targets Genetics platform. Coloc analysis tables between the gene, molecular QTLs, and phenotypes were extracted, with protein QTLs for APOE specifically identified based on blood plasma data from Sun et al.^74^ and Suhre et al.^75^

Similarly for osteoporosis and AD, we utilized the Open Genetics platform to identify shared locus between heel bone mineral density (proxy for osteoporosis) and Family History of AD or AD. To further investigate the locus, we extracted GWAS summary statistics from Jansen et al.^43^ for AD and sex-stratified GWAS summary statistics for heel bone mineral density (HBMD) from Neale’s Lab GWAS round 2, Phenotype Code:3148, based on data from the UK Biobank (www.nealelab.is/uk-biobank/)^76^. We then conducted colocalization analysis using the coloc method described in Giambartolomei et al.^77^, from R package coloc 5.1.0. Summary statistics for MS4A6A cis eQTLs in blood were extracted from eQTLGen^78^, and colocalization analysis was performed between AD, sex-stratified HBMD, and MS4A6A eQTLs on the Locus Region 60050000-60200000 of Chromosome 11. To investigate further associations with the locus, MS4A6A associations with all other phenotypes was extracted from Open Targets Genetics platform with inclusion of a weighted literature evidence association scores.

### Ethical Approval

This study was approved by the Institutional Review Board of University of California San Francisco (IRB #20-32422).

## Data Availability

EHR concepts and identification approaches are described in Methods, and concepts are provided in Supplemental Tables 1 and 2. Phecodes can be downloaded at phewascatalog.org/phecodes_icd10 or phewascatalog.org/phecodes, and mappings between ICD-10 codes and SNOMED can be accessed at www.nlm.nih.gov/healthit/snomedct/us_edition.html. Data for UK Biobank phenotype GWAS can be found at www.nealelab.is/uk-biobank/, and eQTL data can be downloaded from www.eqtlgen.org/. The UCSF EHR database can be accessed to UCSF-affiliated. The SPOKE knowledge network can be accessed at spoke.rbvi.ucsf.edu/, and more details about the network can be found in Morris et al. and mappings to EHR concepts can be found in Nelson et al.

https://www.genetics.opentargets.org/api

https://www.phewascatalog.org/phecodes_icd10

https://www.spoke.rbvi.ucsf.edu/

https://www.eqtlgen.org/

https://www.nealelab.is/uk-biobank/

## Code and Data Availability

EHR concepts and identification approaches are described in Methods, and concepts are provided in Supplemental Tables 1 and 2. Phecodes can be downloaded at phewascatalog.org/phecodes_icd10 or phewascatalog.org/phecodes, and mappings between ICD-10 codes and SNOMED can be accessed at www.nlm.nih.gov/healthit/snomedct/us_edition.html. Data for UK Biobank phenotype GWAS can be found at www.nealelab.is/uk-biobank/, and eQTL data can be downloaded from www.eqtlgen.org/. The UCSF EHR database can be accessed to UCSF-affiliated. The SPOKE knowledge network can be accessed at spoke.rbvi.ucsf.edu/, and more details about the network can be found in Morris et al.^28^ and mappings to EHR concepts can be found in Nelson et al.^27^

## Acknowledgements

Primary support was provided by grant numbers NIA R01AG060393 (AT, SM, SW, TTO, MS). Additional support was provided by the Medical Scientist Training Program T32GM007618 and F30 Fellowship 1F30AG079504-01 (AT) and NSF GRFP 2038436 (JR). SEB holds the Heidrich Family and Friends Endowed Chair of Neurology at UCSF. SEB holds the Distinguished Professorship in Neurology I at UCSF. Dr. Bove is the recipient of a National Multiple Sclerosis Society Harry Weaver Award. She is supported by the NIH, NMSS, NSF, DOD, UCSF Weill Institute for Neurosciences, and by various foundations.Any opinions, findings, and conclusions or recommendations expressed in this material are those of the author(s) and do not necessarily reflect the views of the National Science Foundation. The authors would like to thank everyone in the Sirota Lab for their help and feedback on the analysis approaches and figures. The authors would like to thank the Information Commons and Research Analytics Environment teams for access and support with the UCSF EHR data. The authors would like to thank the SPOKE development team and the UCSF Resource for Biocomputing, Visualization, and Informatics for support in knowledge graph access and maintenance. The authors acknowledge the use of resources developed and supported by the UCSF IT Academic Research Systems and the UCSF Bakar Computational Health Sciences Institute Information Commons groups, and thank all members of these groups for technical support. The authors also thank the Center for Data-driven Insights and Innovation at UC Health (CDI2), for its analytical and technical support related to use of the UC Health Data Warehouse.

