## Supplement Information for "Leveraging Electronic Medical Records and Knowledge Networks to Predict Disease Onset and Gain Biological Insight Into Alzheimer’s Disease"

[4] Weill Institute for Neuroscience. Department of Neurology, University of California, San Francisco, San Francisco, CA

[5] Department of Epidemiology and Biostatistics, University of California, San Francisco, San Francisco, CA

[6] Institute of Developmental and Regenerative Medicine, Department of Paediatrics, University of Oxford, Oxford, OX3 7TY, UK

[7] Department of Psychiatry and Behavioral Sciences, Weill Institute for Neurosciences, University of California, San Francisco, San Francisco, CA, USA

[8] Department of Pediatrics, University of California, San Francisco, San Francisco, CA

### Supplemental Tables and Figures

**Supplemental Figure 1:** Top detailed features and phecodes from the random forest model

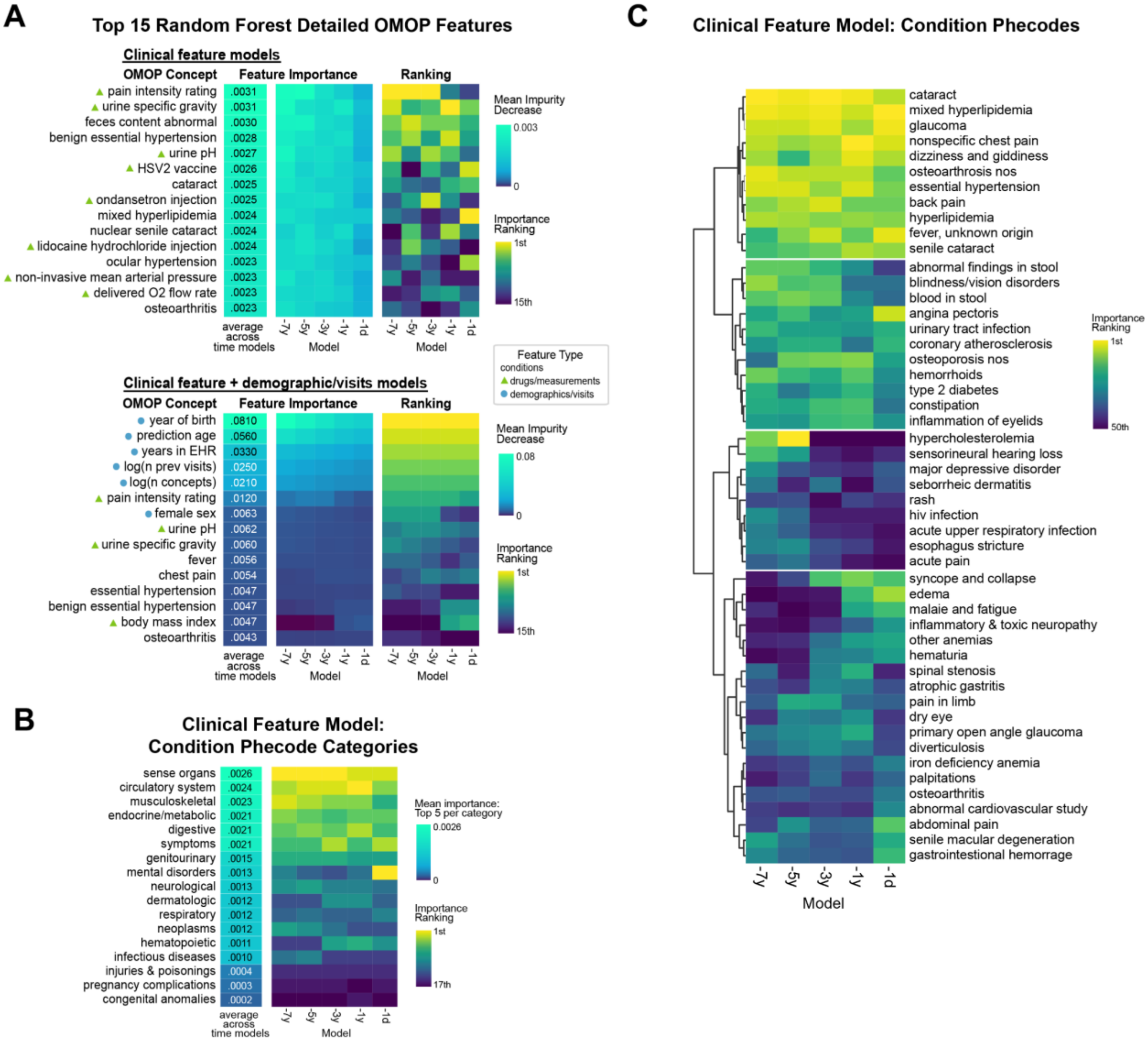


A. Top detailed OMOP clinical features utilized in models for clinical feature only models (top), or clinical features + demographic + visit information models (bottom). Features within the drug/measurement categories are marked with a triangle, while demographic/visit features are marked with a circle.

B. Top phecode categories utilized in models, where importance is determined by the top 5 detailed features within each phecode mapping. The vertical order is based upon the average importance across time models.

C. Top 50 phecodes utilized in time models, clustered based on relative importance across time models.

**Supplemental Figure 2: Comparison of age and visit-related factors between AD, controls, and matched controls.**

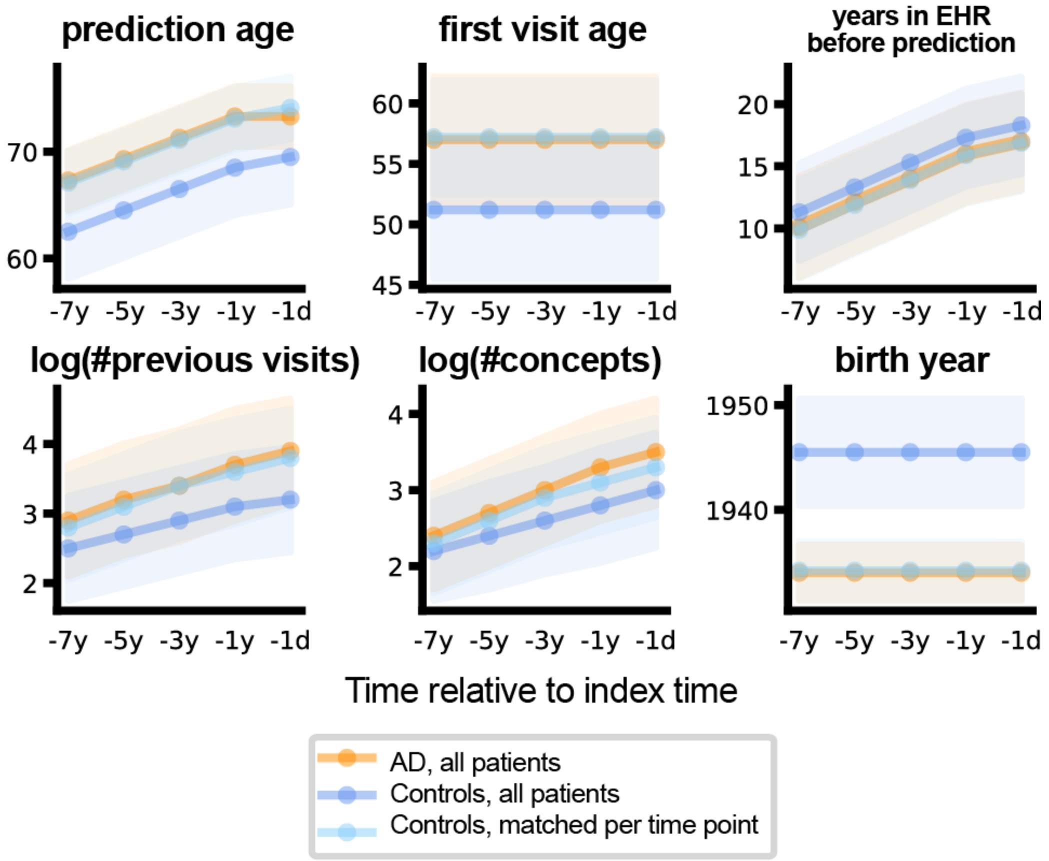


These plots demonstrate the distribution of continuous variables utilized in matching. Orange represents AD patients at each time point. Dark blue represents all controls, while light blue represents controls that have been matched at each time point.

**Supplemental Figure 3:** Sex stratified model performance and features

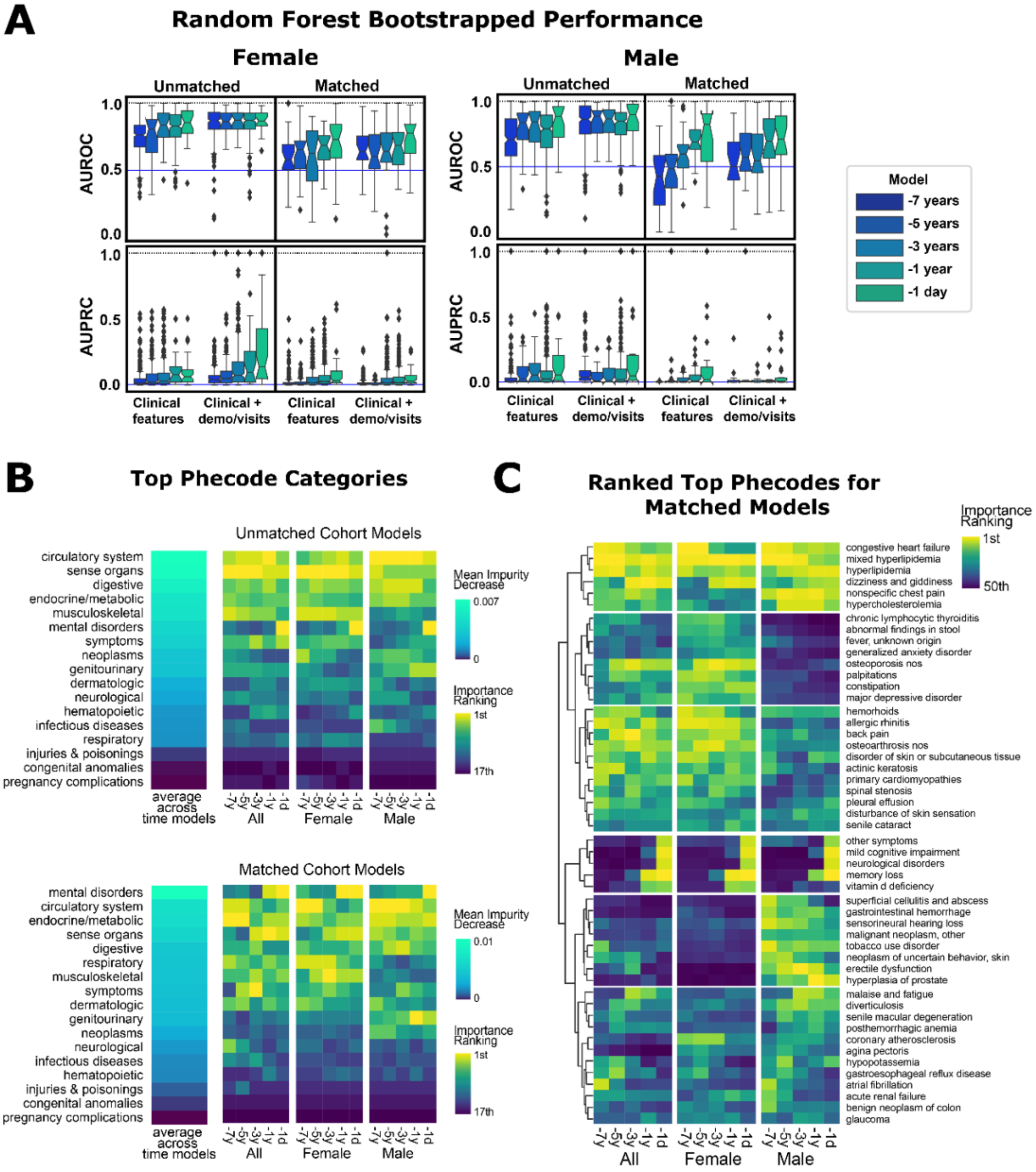


A. The full performance of sex-stratified models are shown. The bootstrapped AUROC/AUPRC is determined by the male or female strata of the initial 30% held-out test set.
B. Top phecode categories are listed by importance for all models, with inclusion of comparison with the general non-stratified model.
C. Top 50 important phecodes clustered by relative importance across time models and across strata.

**Supplemental Figure 4:** UCDDP hyperlipidemia and osteoporosis survival curve numbers and cox proportional hazard model results

**A** Hyperlipidemia → AD survival curve

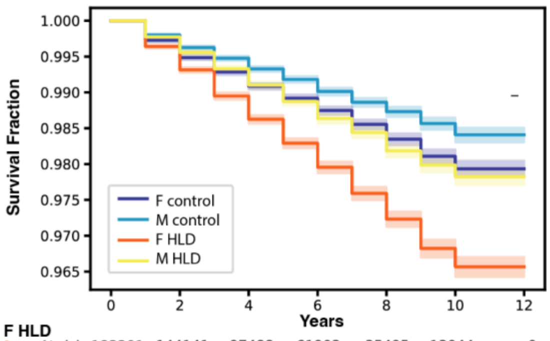


| | 0 | 2 | 4 | 6 | 8 | 10 | 12 |
|---|---|---|---|---|---|---|---|
| **F HLD** | | | | | | | |
| At risk | 183201 | 144141 | 97488 | 61903 | 35495 | 13044 | 0 |
| Censored | 0 | 37863 | 83591 | 118579 | 144581 | 166818 | 179862 |
| Events | 0 | 1197 | 2122 | 2719 | 3125 | 3339 | 3339 |
| **M HLD** | | | | | | | |
| At risk | 178551 | 143424 | 97398 | 61518 | 33611 | 12010 | 0 |
| Censored | 0 | 34374 | 79793 | 115262 | 142925 | 164421 | 176431 |
| Events | 0 | 753 | 1360 | 1771 | 2015 | 2120 | 2120 |
| **F control** | | | | | | | |
| At risk | 185375 | 184420 | 112506 | 58973 | 28016 | 8729 | 0 |
| Censored | 0 | 0 | 71230 | 124432 | 155187 | 174375 | 183104 |
| Events | 0 | 955 | 1639 | 1970 | 2172 | 2271 | 2271 |
| **M control** | | | | | | | |
| At risk | 177265 | 176599 | 111455 | 60617 | 28098 | 9024 | 0 |
| Censored | 0 | 0 | 64658 | 115190 | 147561 | 166560 | 175584 |
| Events | 0 | 666 | 1152 | 1458 | 1606 | 1681 | 1681 |

| Log Rank Test Comparison | test statistic | -log2(p) | pval |
|---|---|---|---|
| all: HLD vs control | 383.32 | 281.13 | 2.36E-85 |
| F: HLD vs control | 308.98 | 227.35 | 3.64E-69 |
| M: HLD vs control | 92.06 | 70.01 | 8.39E-22 |
| F vs M HLD | 255.75 | 188.82 | 1.45E-57 |

**B** Osteoporosis → AD survival curve

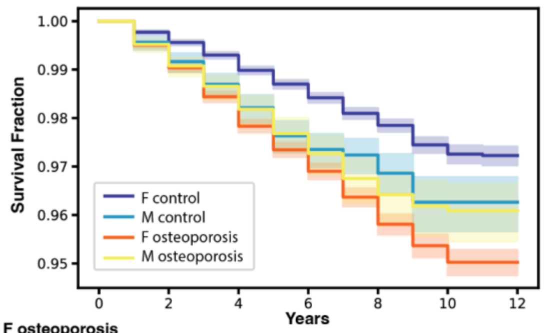


| | 0 | 2 | 4 | 6 | 8 | 10 | 12 |
|---|---|---|---|---|---|---|---|
| **F osteoporosis** | | | | | | | |
| At risk | 58737 | 47341 | 37081 | 25278 | 14093 | 4280 | 0 |
| Censored | 0 | 10854 | 20568 | 32042 | 42975 | 52690 | 56970 |
| Events | 0 | 542 | 1088 | 1417 | 1669 | 1767 | 1767 |
| **M osteoporosis** | | | | | | | |
| At risk | 9597 | 7282 | 5275 | 3208 | 1622 | 474 | 0 |
| Censored | 0 | 2231 | 4177 | 6199 | 7760 | 8903 | 9377 |
| Events | 0 | 84 | 145 | 190 | 215 | 220 | 220 |
| **F control** | | | | | | | |
| At risk | 60141 | 59876 | 59530 | 44003 | 15587 | 3199 | 0 |
| Censored | 0 | 0 | 0 | 15187 | 43391 | 55699 | 58897 |
| Events | 0 | 265 | 611 | 951 | 1163 | 1243 | 1244 |
| **M control** | | | | | | | |
| At risk | 8501 | 8430 | 8349 | 5079 | 1620 | 389 | 0 |
| Censored | 0 | 0 | 0 | 3197 | 6640 | 7861 | 8250 |
| Events | 0 | 71 | 152 | 225 | 241 | 251 | 251 |

| Log Rank Test Comparison | test statistic | -log2(p) | pval |
|---|---|---|---|
| all: osteoporosis vs control | 287.91 | 212.1 | 1.42E-64 |
| F: osteoporosis vs control | 321.39 | 236.33 | 7.22E-72 |
| M: osteoporosis vs control | 0.55 | 1.12 | 4.60E-01 |
| F vs M osteoporosis | 5.14 | 5.42 | 2.34E-02 |

**C** UCDDP: Hyperlipidemia Exposure, AD Diagnosis Outcome

| | No Strata | | | Strata: recruitment age | | |
|---|---|---|---|---|---|---|
| Model | Hazard Ratio | 95% CI | p-value | Hazard Ratio | 95% CI | p-value |
| Unadjusted | **1.53** | [1.47, 1.58] | 2.18E-124 | **1.49** | [1.44, 1.54] | 9.79E-111 |
| demographics adjusted | **1.43** | [1.38, 1.48] | 1.12E-87 | **1.47** | [1.42, 1.53] | 1.43E-104 |
| visit adjusted | **1.32** | [1.26, 1.37] | 9.22E-42 | **1.28** | [1.23, 1.34] | 1.11E-32 |
| visit/demographics adjusted | **1.27** | [1.22, 1.32] | 1.12E-27 | **1.28** | [1.23, 1.33] | 1.51E-31 |

**D** UCDDP: Osteoporosis Exposure, AD Diagnosis Outcome

| | No Strata | | | Strata: recruitment age | | |
|---|---|---|---|---|---|---|
| Model | Hazard Ratio | 95% CI | p-value | Hazard Ratio | 95% CI | p-value |
| Unadjusted | **1.81** | [1.70, 1.92] | 5.20E-82 | **1.71** | [1.61, 1.82] | 7.10E-67 |
| demographics adjusted | **1.61** | [1.52, 1.72] | 1.52E-52 | **1.70** | [1.60, 1.81] | 6.98E-07 |
| visit adjusted | **1.68** | [1.56, 1.80] | 4.34E-47 | **1.59** | [1.48, 1.72] | 4.57E-34 |
| visit/demographics adjusted | **1.57** | [1.45, 1.70] | 7.96E-29 | **1.57** | [1.46, 1.69] | 1.05E-31 |

A. Hyperlipidemia sex-stratified combined Kaplan-Meyer survival curves with counts. Log rank test comparison results are below.
B. Osteoporosis sex-stratified combined Kaplan-Meyer survival curves with counts. Log rank test comparison results are below.

C. Hyperlipidemia exposure cox proportional hazard models for AD as the outcome, shown are the hazard ratios obtained from the exposure coefficient for unadjusted, demographic adjusted (gender, age, race, ethnicity), visit adjusted (first visit age, log(number of visits)), and demographic/visit adjusted. Right group shows computed hazard ratios with stratification by recruitment or starting age (age strata: <55, 55-60, 60-65, 65-70, 70-75, 75-80, >80).
D. Osteoporosis exposure cox proportional hazard models for AD as the outcome, shown are the hazard ratios obtained from the exposure coefficient for unadjusted, demographic adjusted, visit adjusted, and demographic/visit adjusted. Right group shows computed hazard ratios with stratification by recruitment or starting age (age strata: <60, 60-65, 65-70, 70-75, 75-80, >80).

**Supplemental Table 1:** Control exclusion codes.
List of mappings from ICD-10 codes G[123]* to OMOP codes for determining exclusion of Controls. The mapping was generated and manually reviewed to white-list certain codes and approve exclusion of dementia-related codes.

**Supplemental Table 2:** Dementia codes.
List of mappings from Dementia/FTD related condition concepts to SNOMED OMOP mappings and N06D ATC code to RxNorm OMOP mappings for identifying index time 0 for AD patients.

**Supplemental Table 3:** Matching results for every time model
Demographics of matched cohorts (propensity-score matched by demographics and visit-related factors, see Methods) on the training set for hypothesis generation models.

**Supplemental Table 4:** Male and Female demographics and matching result
Demographics of male and female cohorts (combined train and test set). The same patients for train/test set split in the general model are utilized for the sex-stratified models. Matched cohorts on the sex-strata training sets are also shown for hypothesis generation models.

**Supplemental Table 5:** UCDDP AD patient concepts and demographics

Top table shows the specific concepts utilized to identify Alzheimer's Disease as the outcome in the UCDDP database, with breakdown by number of patients per concept. Due to deidentification, only a patient's birth year is known for age estimation.

| **Term** | **# patients** |
|---|---|
| Alzheimer's disease | 20562 |
| Primary degenerative dementia of the Alzheimer type, senile onset | 9327 |
| Primary degenerative dementia of the Alzheimer type, presenile onset | 2530 |

| | | **Overall** |
|---|---|---|
| **n** | | 24389 |
| **estimated_age, mean (SD)** | | 45.6 (23.5) |
| **gender, n (%)** | **FEMALE** | 12915 (53.0) |
| | **MALE** | 11391 (46.7) |
| | **UNKNOWN** | 83 (0.3) |
| **race, n (%)** | **Native** | 78 (0.3) |
| | **Asian** | 2069 (8.5) |
| | **Black** | 1079 (4.4) |
| | **Multirace** | 494 (2.0) |
| | **NHPI** | 108 (0.4) |
| | **Other Race** | 3413 (14.0) |
| | **Unknown** | 6535 (26.8) |
| | **White** | 10613 (43.5) |
| **ethnicity, n (%)** | **Hispanic or Latino** | 3815 (15.6) |
| | **Not Hispanic or Latino** | 13869 (56.9) |
| | **Unknown** | 6705 (27.5) |
| **# visits, mean (SD)** | missing = 3092 | 21.1 (51.8) |

**Supplemental Table 6:** Hyperlipidemia UCDDP concepts and demographics
Top table shows the specific concepts utilized to identify HLD as the exposure in the UCDDP database, with breakdown by number of patients per concept. Due to deidentification, only a patient's birth year is known for age estimation. Recruitment age is utilized as the starting age for survival analysis, with HLD group as the age of HLD diagnosis, and unexposed group as the age of first EHR visit.

| Term | # patients |
|---|---|
| Hyperlipidemia | 702142 |
| Mixed hyperlipidemia | 169316 |

| | | Overall | No HLD | HLD | SMD |
|---|---|---|---|---|---|
| n | | 728578 | 364289 | 364289 | |
| gender, n (%) | FEMALE | 371050 (50.9) | 186259 (51.1) | 184791 (50.7) | 0.037 |
| | MALE | 357255 (49.0) | 177768 (48.8) | 179487 (49.3) | |
| | UNKNOWN | 273 (0.0) | 262 (0.1) | 11 (0.0) | |
| race, n (%) | Native | 3278 (0.4) | 1762 (0.5) | 1516 (0.4) | 0.113 |
| | Asian | 69432 (9.5) | 32466 (8.9) | 36966 (10.1) | |
| | Black | 35072 (4.8) | 16512 (4.5) | 18560 (5.1) | |
| | Multirace | 17486 (2.4) | 7635 (2.1) | 9851 (2.7) | |
| | NHPI | 2972 (0.4) | 1270 (0.3) | 1702 (0.5) | |
| | Other Race | 81646 (11.2) | 44093 (12.1) | 37553 (10.3) | |
| | Unknown | 81062 (11.1) | 44889 (12.3) | 36173 (9.9) | |
| | White | 437630 (60.1) | 215662 (59.2) | 221968 (60.9) | |
| ethnicity, n (%) | H/L | 102163 (14.0) | 53581 (14.7) | 48582 (13.3) | 0.126 |
| | Not H/L | 560067 (76.9) | 271574 (74.5) | 288493 (79.2) | |
| | Unknown | 66348 (9.1) | 39134 (10.7) | 27214 (7.5) | |
| estimated_age, mean (SD) | | 69.7 (10.8) | 69.6 (11.0) | 69.8 (10.7) | 0.012 |
| recruitment_age, mean (SD) | | 63.9 (10.5) | 63.4 (10.5) | 64.3 (10.5) | 0.087 |

**Supplemental Table 7:** Osteoporosis UCDDP concepts and demographics
Top table shows the specific concepts utilized to identify osteoporosis as the exposure in the UCDDP database with inclusion of children concepts, and breakdown by number of patients per concept. Due to deidentification, only a patient's birth year is known for age estimation. Recruitment age is utilized as the starting age for survival analysis, with osteoporosis group as the age of osteoporosis diagnosis, and unexposed group as the age of first EHR visit.

| **Term** | # patients |
|---|---|
| Osteoporosis | 145608 |
| Senile osteoporosis | 30611 |
| Osteoporotic fracture | 7772 |
| Osteoporotic fracture of vertebra | 3987 |
| Localized osteoporosis - Lequesne | 3126 |
| Osteoporotic fracture of femur | 2971 |
| Idiopathic osteoporosis | 1231 |
| Disuse osteoporosis | 309 |
| Osteoporotic fracture of humerus | 186 |
| Osteoporotic fracture of hand | 39 |

| | | **Overall** | **No osteo** | **osteo** | **SMD** |
|---|---|---|---|---|---|
| **n** | | 137880 | 68940 | 68940 | |
| **gender, n (%)** | **FEMALE** | 119637 (86.8) | 60386 (87.6) | 59251 (85.9) | 0.049 |
| | **MALE** | 18241 (13.2) | 8554 (12.4) | 9687 (14.1) | |
| | **UNKNOWN** | 2 (0.0) | | 2 (0.0) | |
| **race, n (%)** | **Native** | 496 (0.4) | 272 (0.4) | 224 (0.3) | 0.134 |
| | **Asian** | 15784 (11.4) | 7364 (10.7) | 8420 (12.2) | |
| | **Black** | 4611 (3.3) | 2546 (3.7) | 2065 (3.0) | |
| | **Multirace** | 3564 (2.6) | 1737 (2.5) | 1827 (2.7) | |
| | **NHPI** | 419 (0.3) | 198 (0.3) | 221 (0.3) | |
| | **Other Race** | 13032 (9.5) | 7427 (10.8) | 5605 (8.1) | |
| | **Unknown** | 13670 (9.9) | 7552 (11.0) | 6118 (8.9) | |
| | **White** | 86304 (62.6) | 41844 (60.7) | 44460 (64.5) | |
| **ethnicity, n (%)** | **H/L** | 15530 (11.3) | 8509 (12.3) | 7021 (10.2) | 0.133 |
| | **Not H/L** | 112474 (81.6) | 54548 (79.1) | 57926 (84.0) | |
| | **Unknown** | 9876 (7.2) | 5883 (8.5) | 3993 (5.8) | |
| **estimated_age, mean (SD)** | | 74.8 (9.2) | 75.2 (9.1) | 74.5 (9.3) | -0.074 |
| **recruitment_age, mean (SD)** | | 68.7 (8.9) | 68.2 (8.7) | 69.2 (9.1) | 0.12 |
